# Linked Patient and Provider Impressions of Outpatient Teleneurology Encounters

**DOI:** 10.1101/2022.12.07.22283213

**Authors:** Justin G. James, Jane Park, Alexandria Oliver, Sharon X. Xie, Andrew Siderowf, Meredith Spindler, Lawrence Wechsler, Thomas F. Tropea

## Abstract

**Background and Objectives:** Teleneurology is common in clinical practice partly due to the SARS CoV-2 pandemic. Impressions about teleneurology from patients and providers alike are generally favorable; some of the reported benefits include ease of access to specialized healthcare, savings of time and money, and similar quality of care as an in-person visit. However, comparisons between patient and provider impressions about the same teleneurology encounter have not been described. Here we describe patient impressions about a teleneurology encounter and evaluate concordance with provider impressions about the same encounter.

**Methods:** Patients and providers at the University of Pennsylvania Hospital Neurology Department were surveyed about their impressions of teleneurology between April 27^th^ and June 16^th^, 2020. A convenience sample of patients, whose providers completed a questionnaire, were contacted by telephone to solicit their impressions the same encounter. Unique questionnaires for patients and providers focused on similar themes, such as adequacy of technology, assessment of history obtained, and overall quality of the visit. Summaries of patient responses are reported with the raw percent agreement between patients and providers for similar questions.

**Results:** One hundred thirty-seven patients completed the survey; 64 (47%) were male and 73 were (53%) female. Sixty-six (47%) patients had a primary diagnosis of PD, 42 (30%) a non-PD movement disorder, and 29 (21%) a non-movement disorder neurological disease. One hundred one (76%) were established patient visits and 36 (26%) were new patient visits. Provider responses from 8 different physicians were included. The majority of patients responded that the ease of joining their visit, their comfort engaging with their physicians during their visit, understanding their plan of care after their visit, and the quality of care from their teleneurology visit were satisfactory. Patients and providers agreed about their impressions of the quality of the history obtained (87% agreement), patient-provider relationship (88% agreement), and overall quality of their experience (70% agreement).

**Discussion:** Patients had favorable impressions about their clinical experience with teleneurology and expressed an interest in incorporating telemedicine visits into their ongoing care. Patients and providers were highly concordant for the history obtained, patient-provider relationship, and overall quality.

## Introduction

Teleneurology, or the delivery of neurological care via telephone or videoconference, was widely adopted during the severe acute respiratory syndrome coronavirus 2 (SARS CoV-2) pandemic. This allowed for continuity in patient care despite the challenges posed by technology access and limited evidence of best practices.^1,2^ Prior to the pandemic, research on stroke care via telemedicine demonstrated improved access and quality of care, leading to telestroke to become mainstream practice.^3,4^ These contributions led to telemedicine being applied to other neurological conditions, such as Parkinson’s Disease (PD), where survey studies conveyed positive impressions from patients and providers in the outpatient setting.^5,6^ The expansion of teleneurology across all neurology sub-specialties in the United States and abroad during the pandemic allowed for observations about physician and patient preferences, safety, and shortfalls of teleneurology that will be informative beyond the pandemic.^7^

As key stakeholders, patients and their medical care providers have valuable insight into the teleneurology experience that can inform clinical care and future research studies. Previous reports of provider impressions of outpatient teleneurology have demonstrated that the time required for visit, ability to connect with patients, and quality of care delivered during the visit was perceived similar or equivalent to that of an in-person visit.^8,9^ Similarly, patient experiences with teleneurology in the outpatient setting have conveyed high satisfaction with the quality of care provided and the convenience of telemedicine, such as time and money saved.^10,11,12^

However, few to no studies have reported the experiences of patients and providers from the same telemedicine encounter to evaluate if there is agreement in their impressions. Examining linked patient and provider encounters allow us to understand the agreement and disagreement in clinical evaluations between patients and providers about the same completed teleneurology. In return, this method of report can justify policies supporting the expansion of telemedicine to improve access to neurological care and inform clinicians how telemedicine should be integrated into outpatient care.

Here, we present comparisons between patient and provider impressions about the same, distinct teleneurology encounter at the Pennsylvania Hospital Department of Neurology. Our hypothesis-generating, quality improvement observations aim to describe patient impressions about their teleneurology experience and to evaluate the level of concordance between patient and provider opinions of teleneurology.

## Methods

### Survey Design

A novel questionnaire was developed by authors M.S., A.S., J.P., L.W., and T.F.T, to be administered to patients after their teleneurology encounter. The patient survey questions were designed to solicit feedback on clinical impressions, technology adequacy, and perceived benefits from their teleneurology experience with the goal of quality improvement. Questions on this survey were similar to a provider questionnaire we previously reported.^8^ However, wording on the similar questions between the surveys were adjusted to be administered to patients. See **Table 1** for all questions and possible responses on the patient questionnaire.

**Table 1.**
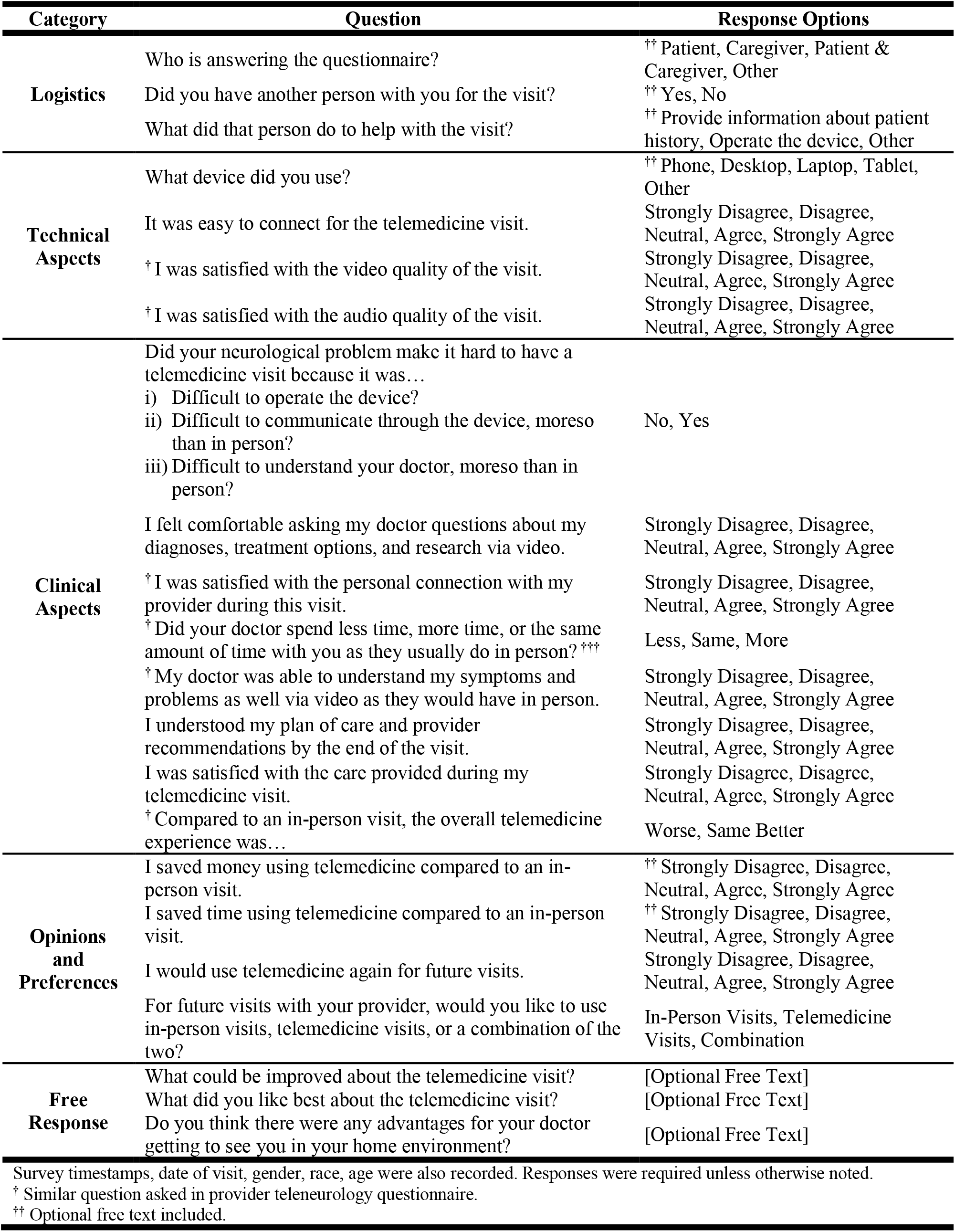
Teleneurology Patient Questionnaire.

After 73 patient responses were collected, five questions were updated or removed at the discretion of the authors to improve comprehension and reporting by the patients. Three questions asking “*Would you have preferred to go to a local clinic where someone could have set up the technology for you (assuming COVID was not a risk)?*”, “*Would you be more likely to participate in a support group if it were available via video versus in-person (so you didn’t have to drive to it)?*”, and “*If your primary doctor referred you to a specialist in the city, would you be more likely to see them if they could do a video visit instead of an in-person visit?*” were removed because of poor wording resulting in confusion among the respondents. One question asking, “*I prefer in-person visits to telemedicine visits*.” was modified to ask, “*For future visits with your provider, would you like to use in-person visits, telemedicine visits, or a combination of the two?*”. One question asking to report the amount of time required during the teleneurology visit compared to an in-person visit was only asked to returning patients. Data reported are based on the available data.

### Survey Distribution and Collection

A convenience sample of patients who completed an outpatient audiovisual teleneurology visit at the Pennsylvania Hospital Department of Neurology between May 5 to June 24, 2020 were contacted within 14 days to complete the survey over the telephone. A single research coordinator (J.P.) administered all surveys and recorded data directly into the patient questionnaire created on REDCap, as shown in ***Supplementary Table 1***.^13^ Patients were selected and contacted about participation only if their provider completed a teleneurology questionnaire from the same audiovisual encounter, as previously reported.^8^ Every patient survey collected had a linked provider survey pertaining to the same encounter. Survey responses of patients collected >14 days after their visit and audio/telephone-only telemedicine encounters were excluded from the analysis (13, 8.66%).

### Statistical Analyses

Descriptive statistics are reported for patient demographics. Omnibus Pearson χ^2^ analyses were conducted to compare the frequency of responses with patient and visit characteristics. Where similar questions were asked of patients and providers, the raw percent of similar responses was used to measure the level of agreement between groups.^14^ Due to slight differences in the wording and response options, the possible response options for history obtained, patient-provider relationship, and overall quality of telemedicine visit were adjusted to allow for this comparison. Multiple author perspectives were included (J.G.J., T.F.T, M.S., L.W.) when comparing the patient and provider response questions and options to proceed with concordance analysis.

Data were analyzed using R Studio.^15^ Figures were created in GraphPad Prism Version 9 (GraphPad Software; San Diego, CA; www.graphpad.com). Alpha was set at 0.05 without correction for multiple testing.

### Standard Protocol, Approvals, Registrations, and Patient Consents

This project was reviewed and determined to qualify as quality improvement by the University of Pennsylvania’s Institutional Review Board. Therefore, informed consent was not obtained. Revised Standards for Quality Improvement Reporting Excellence (SQUIRE 2.0) reporting guidelines were followed.^16^

## Results

### Cohort Summary

Surveys were completed by providers and patients between April 27 to June 16, 2020, and May 5 to June 24, 2020, respectively. A total of 1255 (78.63%) videoconference visits were conducted during this time at Pennsylvania Hospital, while 1596 total outpatient visits (341, 21.37% in-person) were conducted. One hundred and fifty patients who completed audiovisual teleneurology visits were contacted and completed the survey, and 13 (8.66%) entries were removed from the analysis. See **Figure 1** for a summary of included participants.

**Figure 1:**
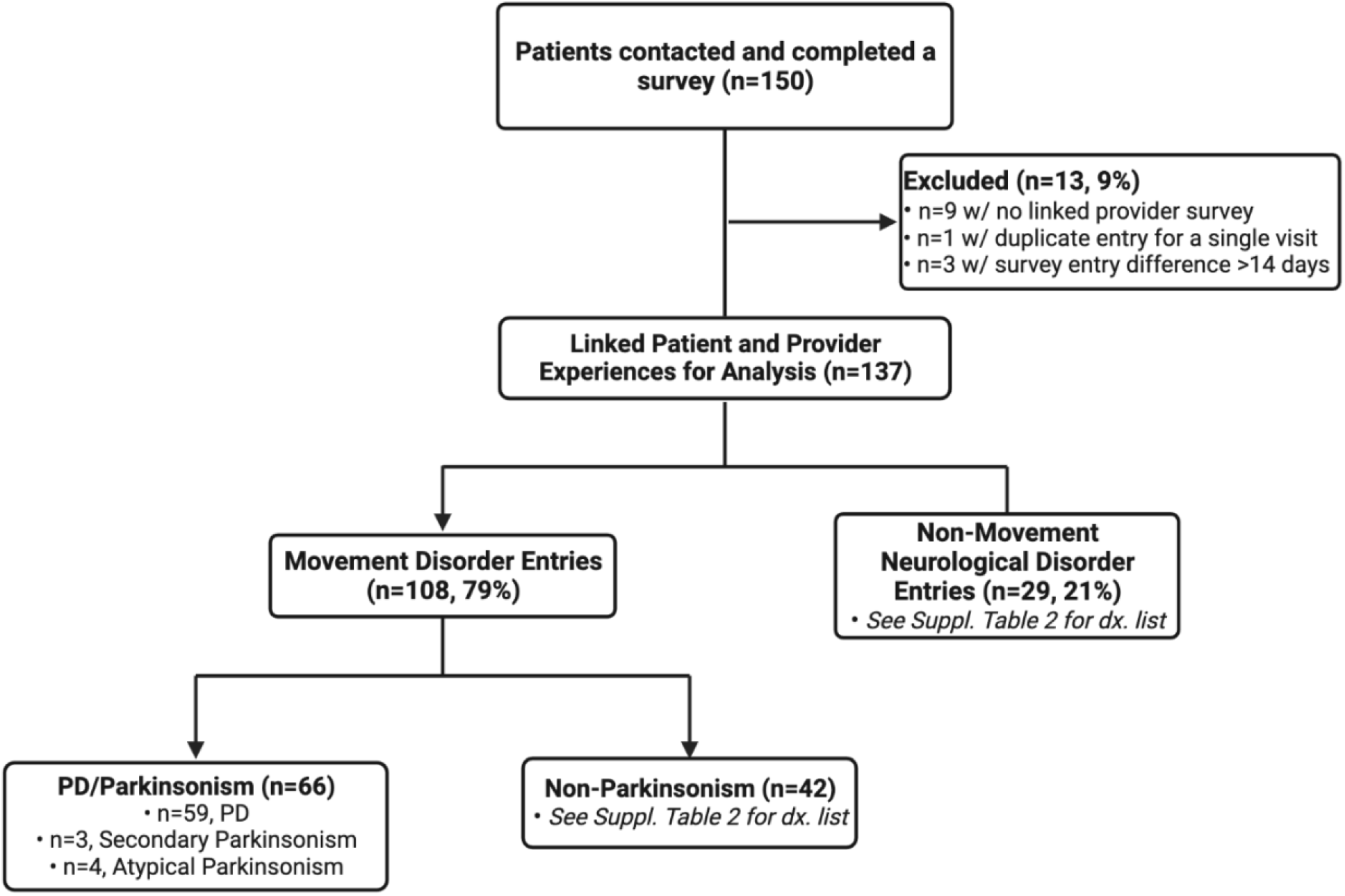
Flowchart of Patient Cohort Included in the Study. Flowchart illustrating the patient cohort contacted and included in completing the teleneurology questionnaires. Abbreviations: *PD = Parkinson’s Disease*.

### Visit Summary

The majority of teleneurology visits were completed through BlueJeans (132, 96.35%), the preferred video conference platform for telemedicine visits by UPHS; however, encounters recorded were also conducted through FaceTime (3, 2.19%) or Doximity (2, 1.46%). The most common devices used by patients for their visits included smartphones (51, 37.23%) and laptops (50, 36.50%), while tablets (24, 17.52%) and desktop computers (12, 8.76%) were less frequently chosen.

Patients assessed the quality of the video to be satisfactory for 95% of encounters and the audio to be satisfactory in 91% of encounters. Moreover, for the majority of patients, their neurological problem did not make it difficult to operate the device (115, 83.94%) or make it more difficult, compared to in-person visit, to communicate (123, 89.78%) or understand their doctor (130, 94.89%) through their device during the telemedicine visit.

### Provider and Patient Characteristics

Patient respondent demographic and visit summaries are found in **Table 2**. Of the 137 unique patient encounters included, questionnaires were completed by patients (116, 84.67%), caregivers (15, 10.95%), or together by patient and caregiver (6, 4.38%). The age of patients ranged from 18 to 86, with a mean age of 58.5 (SD ± 17.87). The primary diagnoses of patients at the time of visits were recorded from their electronic medical record and then categorized as follows: PD/Parkinsonism Movement Disorder (66, 48.18%), Non-PD/Parkinsonism Movement Disorder (42, 30.66%), and Non-Movement Neurological Disorder (29, 21.17%). The diagnoses included in these categories are provided in ***Supplementary Table 2***. The median time from visit completion to survey completion was 8 days (Interquartile Range [IQR] 5-11 days), with 14 days (2, 1.46%) being the longest.

**Table 2.** Patient Cohort Characteristics.

|  | Whole Cohort | PD/Parkinsonism Movement Disorder | Non-PD/Parkinsonism Movement Disorder | Non-Movement Neurological Disorders |
| --- | --- | --- | --- | --- |
|  | 137 | 66 (48) | 42 (31) | 29 (21) |
| **Gender** |  |  |  |  |
| Male | 64 (47) | 38 (58) | 18 (43) | 8 (28) |
| Female | 73 (53) | 28 (42) | 24 (57) | 21 (72) |
| **Age Groups** |  |  |  |  |
| 18-29 | 5 (4) | 0 (0) | 1 (2) | 4 (14) |
| 30-49 | 18 (13) | 1 (2) | 9 (21) | 8 (28) |
| 50-69 | 68 (50) | 37 (56) | 20 (48) | 11 (38) |
| 70-89 | 46 (34) | 28 (42) | 12 (29) | 6 (21) |
| **Race** |  |  |  |  |
| White | 103 (75) | 55 (83) | 29 (69) | 19 (66) |
| Black | 16 (12) | 4 (6) | 5 (12) | 7 (24) |
| Asian | 1 (1) | 1 (2) | 0 (0) | 0 (0) |
| Hispanic Latino | 1 (1) | 1 (2) | 0 (0) | 0 (0) |
| Other | 1 (1) | 0 (0) | 0 (0) | 1 (3) |
| Unknown | 15 (11) | 5 (8) | 8 (19) | 2 (7) |
| **Distance (miles) from Clinic** |  |  |  |  |
| 0-19 | 83 (61) | 36 (55) | 27 (64) | 20 (69) |
| 20-39 | 31 (23) | 14 (21) | 10 (23) | 7 (24) |
| > 40 | 23 (17) | 16 (24) | 5 (12) | 2 (7) |
| **Visit Type Status** |  |  |  |  |
| Existing/Follow-Up | 101 (76) | 54 (82) | 30 (71) | 17 (59) |
| New | 36 (26) | 12 (18) | 12 (29) | 12 (41) |
| **Device Used** |  |  |  |  |
| Phone | 51 (37) | 24 (36) | 19 (45) | 8 (28) |
| Laptop | 50 (36) | 24 (36) | 13 (31) | 13 (45) |
| Tablet | 24 (18) | 15 (23) | 2 (5) | 7 (24) |
| Desktop | 12 (9) | 3 (5) | 8 (19) | 1 (3) |
| Data reported as N (%) of total cohort. |  |  |  |  |
| Abbreviations: PD, Parkinson's Disease. |  |  |  |  |

The provider respondents in this analysis are a subset of the full provider survey completed as previously reported.^8^ Eight physician respondents were included in this analysis: 3 (37.50%) were male and 5 (62.50%) were female. The median years of provider experience (calculated as the time in years since their medical degree was received) was 14 years (range 9-28 years). The median number of surveys completed per provider was 18 (interquartile range [IQR] 9.75-22). Two different departmental divisions were represented in the provider cohort: Movement Disorders (6, 75.00%) and General Neurology (2, 25.00%). The Movement Disorder providers completed 108 (79.00%) surveys while the General Neurology providers completed 29 (21.00%). Ninety-four (68.61%) providers completed the survey response on the same day as the encounter, while 43 (31.39%) completed it within 3 days.

### Patient Impressions of Teleneurology

Patients’ impressions about the teleneurology experience are summarized in **Figure 2 and Supplementary Table 3**. Patient gender, age group, distance from neurology clinic, visit type, device used, and primary diagnosis category did not affect the frequency of patients’ responses in exploratory analyses regarding patient impressions of the history obtained, patient-provider relationship, amount of time required, or the overall quality of the teleneurology visit (*P > 0*.*05*). See ***Supplementary Tables 4-7*** for frequency of responses per category explored in analysis.

**Figure 2:**
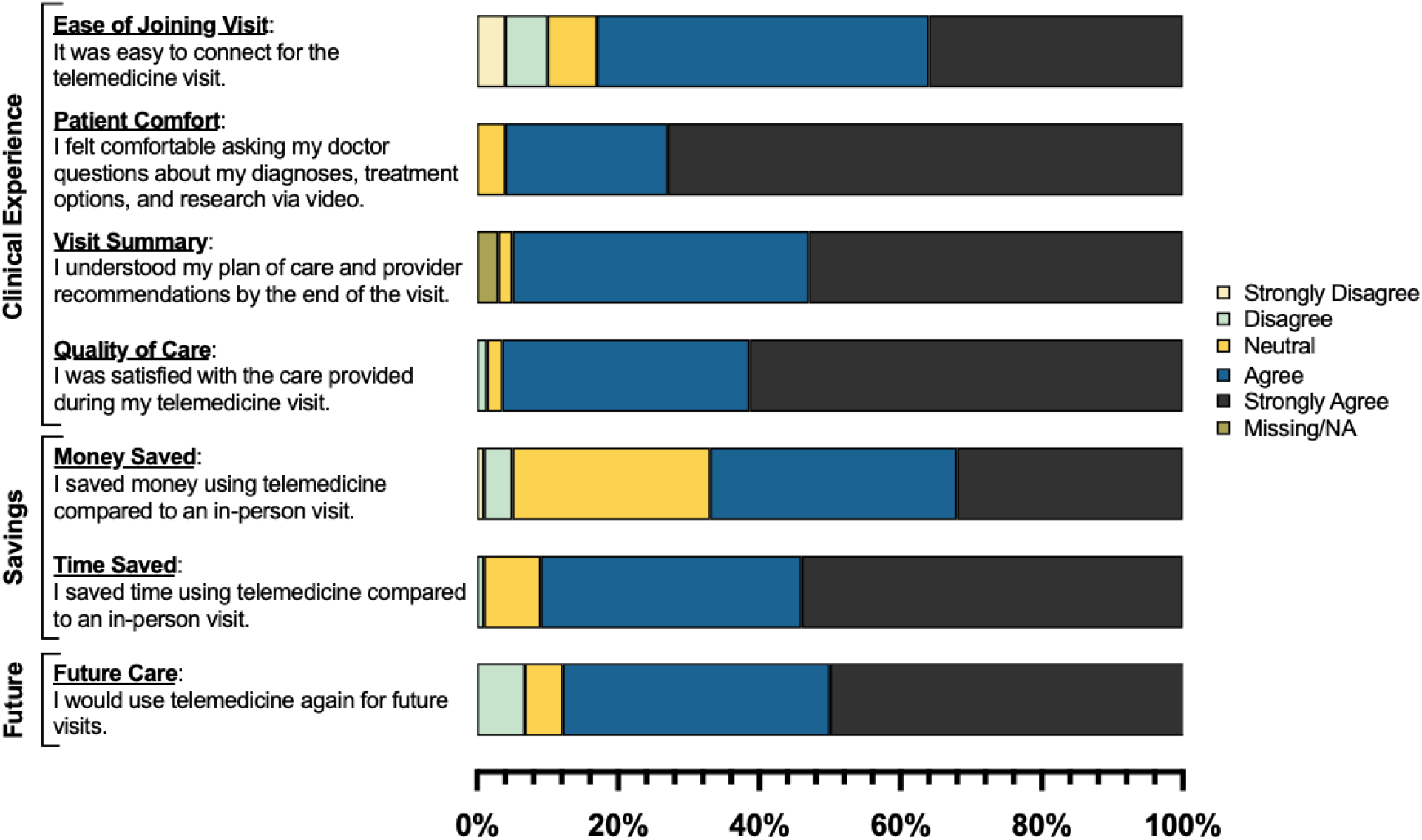
Patient Impressions of Teleneurology Experience. Patient impressions evaluating the clinical aspects and personal benefits of their teleneurology experience. The percent of patients responding to each response category for each question is illustrated in the colored, horizontal bars.

### Linked Patient-Provider Teleneurology Impressions

A summary of linked impressions between patients and providers from the same teleneurology encounter are summarized in **Figure 3**.

**Figure 3:**
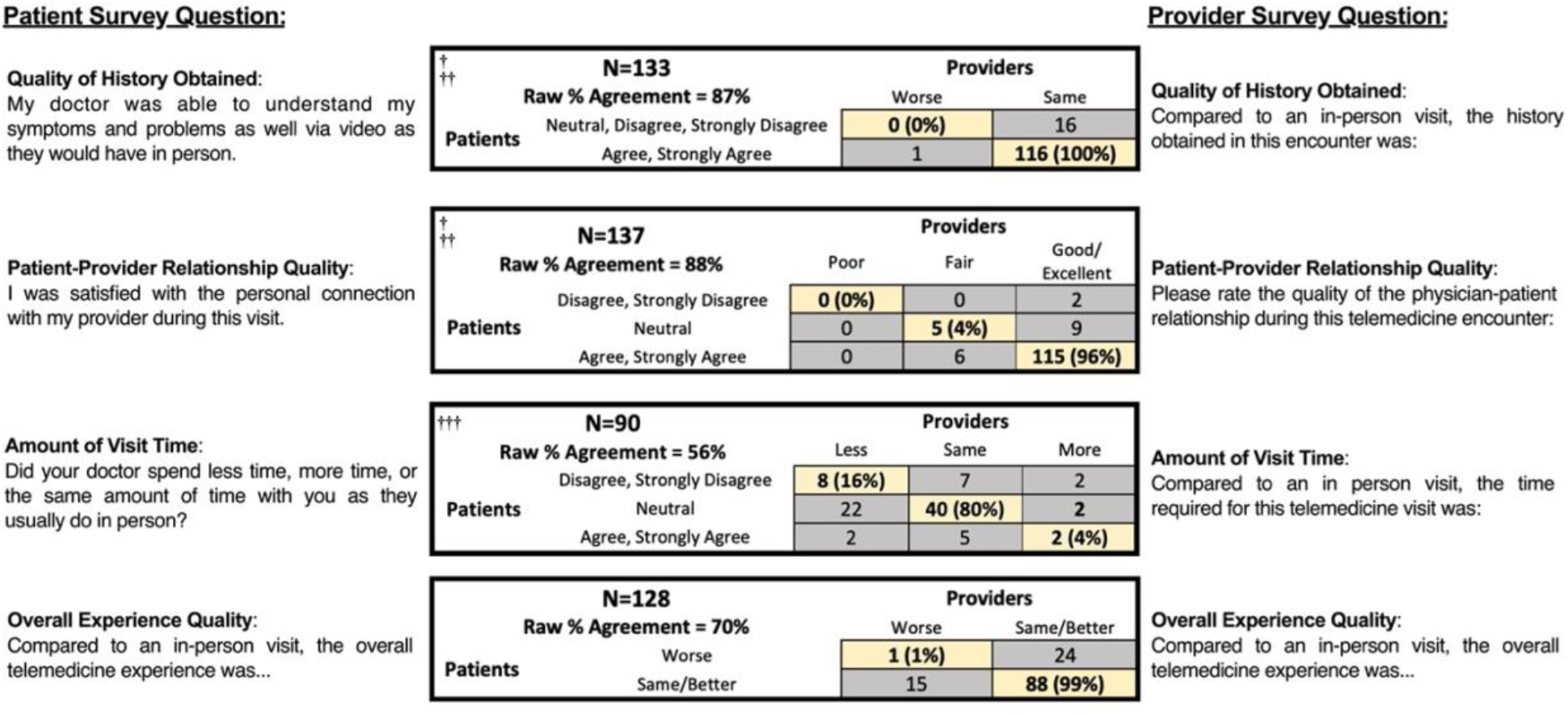
Agreement Between Patient and Provider Clinical Impressions of Teleneurology. The raw percent agreement comparing the teleneurology clinical impressions of patients and providers from the same visit encounter. Raw percent agreement reported as percent agreement of the entire cohort responding to the corresponding question. Highlighted diagonals illustrate the frequency and percentage of paired patient-provider responses that respond the same in each category. ^**†**^ Provider responses and corresponding patient response with “Better” were removed for concordance analysis. ^**††**^ Response options were modified for concordance analysis. ^**†††**^ Established patient responses only included for concordance analysis.

To assess the quality of the history obtained, providers were asked, “*Compared to an in-person visit, the history obtained in this encounter was…*” with the options Worse, Same, or Better, while patients were asked, “*My doctor was able to understand my symptoms and problems as well via video as they would have in person*,” with a 5-point Likert-scale, ranging from Strongly Disagree to Strongly Agree. The response options were adjusted to compare patient and provider impressions. A provider responding Worse was equated to a Neutral, Disagree, or Strongly Disagree response by the patient, and a provider responding Same was aligned with Agree or Strongly Agree by the patient. The providers responding with Better and corresponding linked patient responses were excluded (N=4, 2.92%) for better concordance between response options, leaving 133 (97.08%) paired patient-provider responses included for analysis. One hundred and seventeen (87.97%) patients and 132 (99.25%) providers responded that the history obtained was the same, while 16 (12.03%) patients and 1 (0.75%) provider reported that it was worse. The raw percent agreement showed 87% (116) of linked patient-provider encounters conveying agreement. Of the patient-provider responses in agreement, 100% (116) reported that the quality of the history obtained in their teleneurology visit was the Same as an in-person visit.

In assessing the quality of the patient-provider relationship, providers were asked, “*Please rate the quality of the physician-patient relationship during this telemedicine encounter…*” with the options Poor, Fair, Good, or Excellent, while patients were asked, “*I was satisfied with the personal connection with my provider during this visit*,” with a 5-point Likert-scale, ranging from Strongly Disagree to Strongly Agree. The response options were again adjusted to compare patient and provider impressions. Patients responding Disagree or Strongly Disagree were aligned with a provider response of Poor, a Neutral response by the patients were aligned with a provider responding Fair, and patients responding Agree or Strongly Agree were aligned with a provider responding Good or Excellent. No patient or provider responses were removed for analysis of this question, including all 137 (100%) linked patient-provider responses. One hundred and twenty-six (91.97%) patients and 131 (95.62%) providers reported that the quality of the relationship was Good/Excellent, while 9 (6.57%) patients and 6 (4.38%) providers reported it was Neutral/Fair and 2 (1.46%) patients and 0 (0.00%) providers reported it was Poor. The raw percent agreement showed 120 (88%) paired patient-provider responses agreed. Ninety-six percent (115) of the patient-provider responses in agreement reported that the quality of the patient-provider relationship was Good/Excellent and 4% (5) responded it was Neutral/Fair.

To compare the time spent in this teleneurology visit, providers were asked, “*Compared to an in person visit, the time required for this telemedicine visit was…*” with the options Less, Same, or More. Only follow-up or existing patients were asked the question, “Did your doctor spend less time, more time, or the same amount of time with you as they usually do in person?” with the same options Less, Same, and More. Forty-seven (34.30%) new patient visit patient-provider surveys responses were excluded; 90 responses (65.69%) were included in this analysis. Sixty-four (71.11%) patients and 52 (57.78%) providers responded that the amount of time spent during the telemedicine visit was the Same as an in-person visit, while 17 (18.89%) patients and 32 (35.56%) providers reported it was Less, and 9 (10.00%) patients and 6 (6.67%) providers sharing it was More. The raw percent agreement showed that 56% (50) of patients-provider responses from the same encounter agreed. Of the patient-provider responses in agreement, 80% (40) reported that it required the same amount of time, 16% (8) said it required less time, and 4% (2) responded it took more time.

The overall quality of the telemedicine experience compared to an in-person visit was evaluated by patients and providers being asked, “*Compared to an in person visit, the overall telemedicine experience was…*” and responding with Worse, Same, or Better. Responses with Same or Better were combined for improved interpretation and comparison. Nine (6.57%) patient-provider responses were excluded from the analysis due to missing responses. One hundred and three (80.47%) patients and 112 (87.50%) providers responded that the overall experience was the Same or Better, while 25 (19.53%) patients and 16 (12.50%) providers reporting it was Worse. The raw percent agreement showed that 70% (89) of linked patient-provider responses agreed. Of the patient-provider responses in agreement, 99% (115) reported that the overall teleneurology experience was the “Same/Better” and 1% (1) pair responded it was “Worse” compared to an in-person visit.

## Discussion

In this study, we report on patient impressions of outpatient teleneurology encounters during the SARS CoV-2 pandemic. We showed that patients had highly favorable impressions on several aspects of their clinical experience. Moreover, patients overwhelmingly agreed that using telemedicine allowed them to save money and time compared to an in-person visit. When asked if they would use telemedicine again, the majority of patients responded that they would and that they would prefer a hybrid model of in-person and telemedicine for future care versus an in-person or telemedicine visit alone. Additionally, we leveraged data collected and previously reported on regarding providers impressions from the same encounter to assess concordance between patient and provider.^8^ The rates of agreement between patients and providers were high for the quality of the history obtained (87% raw agreement), the patient-provider relationship (88% raw agreement), and the overall experience of the visit (70% raw agreement). The high concordance of positive impressions of teleneurology encounters is evidence that teleneurology is effective and palatable to both providers and patients alike. The results of this study should encourage physicians to integrate teleneurology as well as endorse policies that expand coverage and use of teleneurology in the outpatient setting.

Previous reports assessing telemedicine evaluated the experiences and impressions from either patients or providers, but not both. Prior reports on outpatient providers’ teleneurology have demonstrated positive impressions about the clinical experience, relationship and connection with their patients, and delivery of care.^8,9^ On the other hand, prior reports on patient impressions about teleneurology have shown general satisfaction with the experience or convenience reported as time and money saved.^10,11,12^ We were unable to find any reports or surveys that gather impressions from patients evaluating clinical aspects of their teleneurology visit, such as their provider understanding their health history, their engagement with their clinician regarding treatment plans for their diagnosis, or their understanding of their plan of care after their visit. Capturing this information from patients is key to understanding the value of teleneurology beyond just patient satisfaction and convenience.

It also helps to identify where teleneurology is insufficient. For example, patients were queried on what about their teleneurology experiences could be improved, and the most frequent comments made by patients concerned communication prior regarding what they would be expected to do during the teleneurology visit and regarding any delays as to when their provider will log on. For instance, as one patient describes their response, “*[I] would prefer if there was an emphasis and explanation that there needs to be room for walking (during assessment) and that, as a Parkinson’s patient, laptop should be preferred*.” Other comments provided insight on how the infrastructure of telemedicine consultations can be improved, such as implementing a virtual waiting room to communicate any delays as to when the provider will log on and adding live closed captioning during the virtual consultations for further clarity during conversations with providers.

Patients were also asked to summarize what their plan of care was after their visit. While the majority of existing patients reported a modification in their medications (49, 48.51%), several new patients (11, 29.73%) were asked by their provider to complete further scans or tests, such as brain imaging, genetic testing, or blood work. When comparing these patient reports to their provider’s response assessing whether there were elements of the examination that might have changed their assessment and plan if performed in-person, we found that 64% (7) providers of new patients who requested further testing responded yes to this question. Specifically, the providers noted that they would have benefited from performing hands-on motor, sensory, reflexive, or gait assessments. This infers that new patients may benefit more from an in-person consultation over teleneurology in obtaining a comprehensive examination and direction of care.

Teleneurology only allows for a limited neurological examination that may affect the plan of care and treatment decisions. This may affect diagnosis groups differently. Movement Disorders is the largest subspeciality at Pennsylvania Hospital and, therefore, 48% of the participants in this report have PD or parkinsonism. We included patients with PD/parkinsonism alongside non-PD/parkinsonism movement disorders and non-movement neurological disorders to evaluate how different disease group may affect the teleneurology impressions. **Supplementary Tables 4-7** illustrate that these categories of primary diagnosis did not have any association in the frequency of patients’ impressions to of the history obtained, patient-provider relationship, amount of time required, or the overall quality of the teleneurology visit. Our results demonstrate that the lack of equivalent motor examination to an in-person visit did not impact the patients’ satisfaction or clinical impressions with their teleneurology consultation.

The strengths of our study should be noted. First, our study is unique in that we compare patient and provider impressions regarding telemedicine from the same outpatient encounter. Furthermore, the breadth of topics covered in the patient questionnaire included information assessing aspects of the clinical experience, components of the patient-provider interaction, and advantages and challenges of telemedicine in neurological care. Capturing information about cost, time saved, and effectiveness of teleneurology for patient care are useful preliminary data to justify further study and expansion of telemedicine. There are several limitations that should be acknowledged within our survey. First, among the 137 provider questionnaires completed, 23% were completed by a single provider (M.S.). This may result in sampling bias with more favorable views of teleneurology being surveyed more frequently. Second, selection bias may have been introduced due to our patient respondents being selected and contacted in a non-random manner. The lack of random sampling or other methods to reduce bias in selection may lead to our cohort sample not being representative of the population. This was a convenience sample as part of a quality improvement project. Third, comparisons between patient and provider responses could not be completely aligned due to the differences in ways that the patients and providers were surveyed. Questions compared concerning the quality of the history obtained or the patient-provider relationship were asked differently to patients and providers intentionally to allow patients and providers to better understand the questions being asked. Nonetheless, to address this, multiple author perspectives were included (J.G.J., T.F.T, M.S., L.W.) when comparing the patient and provider response questions and options. Finally, the small sample size in this quality improvement, exploratory study resulted in low response frequencies in some categories. The Cohen’s Weighted Kappa statistic, a commonly used measure of agreement, underestimates the true agreement with low response frequencies. Additionally, raters were physicians and patients who were untrained, meaning this was their first experience rating their teleneurology visit, which increases risk of guessing on responses. In this scenario of low response rates and untrained raters, percent agreement is the most appropriate statistic to detect agreement between raters.^17^

Here we report the first comparison of patient and provider impressions regarding teleneurology from the same outpatient encounter. Beyond our results illustrating patients’ overall satisfaction with their telemedicine experience, we demonstrate that their impressions are highly concordant with their providers’ opinions of the quality of the history of the patient obtained, the patient-provider relationship, and the overall experience of the visit. Further analysis to identifying best uses of teleneurology should be explored to identify patient, provider, and visit characteristics associated with successful encounters. Although teleneurology may not be best for all outpatient scenarios, we think these observations will shed light on optimal use of teleneurology as part of the treatment paradigm.

## Data Availability

Data will be made available by requesting access from the corresponding author, Thomas F. Tropea.

## Acknowledgments

We thank our patients and all of the neurology providers in the Department of Neurology for their generous time in completing questions.

## Data Availability Statement

Data will be made available by requesting access from the corresponding author.

## Funding Information

Funding was provided by the Department of Neurology, Perelman School of Medicine, University of Pennsylvania. Dr Tropea is additionally funded by the NINDS (K23-NS114167-01A1).

## Notes

### Competing Interest Statement

The authors have declared no competing interest.

### Author Declarations

This project was reviewed and determined to qualify as quality improvement by the University of Pennsylvania's Institutional Review Board.

