## Supplemental Data for "Linked Patient and Provider Impressions of Outpatient Teleneurology Encounters"

### Supplementary Tables/Figures

#### Supplementary Table 1. Patient Teleneurology Questionnaire as Completed via the REDCap Survey Tool

##### Information

| | |
|---|---|
| Subject | ______________________________ |
| MRN | ______________________________ |
| Gender | ◯ Male<br>◯ Female |
| Age | ______________________________ |
| Race | ______________________________ |

##### Questionnaire

| | |
|---|---|
| Who is answering the questionnaire? | ◯ Patient<br>◯ Caregiver<br>◯ Patient & Caregiver<br>◯ Other |
| If other, | ______________________________ |
| What did you like best about the telemedicine visit? | ______________________________ |
| Do you think there were any advantages for your doctor getting to see you in your home environment? | ______________________________ |
| What could be improved about the telemedicine visit? | ______________________________ |
| What device did you use? | ◯ Phone<br>◯ Desktop<br>◯ Laptop<br>◯ Tablet<br>◯ Other |
| If other, | ______________________________ |
| What would you have done for this visit if telemedicine was not available? | ◯ Phone Call<br>◯ Seen a different doctor in person<br>◯ Not seen a doctor |
| FOLLOW-UP PATIENTS ONLY:<br>Did your doctor spend less time, more time, or the same amount of time with you as they usually do in person? | ◯ Less time<br>◯ More time<br>◯ Same amount of time |
| Did you have another person with you for the visit? | ◯ Yes<br>◯ No |
| If yes, who was it? | ◯ Husband/Wife<br>◯ Adult Child<br>◯ Friend<br>◯ Professional Caregiver<br>◯ Other |
| If other, | ______________________________ |

| | |
|---|---|
| I saved time using telemedicine compared to an in-person visit. | ◯ Strongly Agree ◯ Agree ◯ Neutral ◯ Disagree ◯ Strongly Disagree |
| If agree or strongly agree, about how much time did you save? | ________________________ |
| I felt comfortable asking my doctor questions about my diagnoses, treatment options, and research via video. | ◯ Strongly Agree ◯ Agree ◯ Neutral ◯ Disagree ◯ Strongly Disagree |
| My doctor was able to understand my symptoms and problems as well via video as they would have in person. | ◯ Strongly Agree ◯ Agree ◯ Neutral ◯ Disagree ◯ Strongly Disagree |
| I understood my plan of care and provider recommendations by the end of the visit. | ◯ Strongly Agree ◯ Agree ◯ Neutral ◯ Disagree ◯ Strongly Disagree |
| What was the plan after the visit, or the doctor's recommendations at the end of the visit, as you recall it? | ________________________ |
| I was satisfied with the care provided during my telemedicine visit. | ◯ Strongly Agree<br>◯ Agree<br>◯ Neutral<br>◯ Disagree<br>◯ Strongly Disagree |
| For future visits with your provider, would you like to use in-person visits, telemedicine visits, or a combination of the two? | ◯ In-person Visits<br>◯ Telemedicine Visits<br>◯ Combination |
| Would you have preferred to go to a local clinic where someone could have set up the technology for you (assuming corona virus was not a risk)? | ◯ Yes<br>◯ No<br>◯ Unsure |
| Would you be more likely to participate in a support group if it were available via video versus in-person (so you didn't have to drive to it)? | ◯ Yes<br>◯ No<br>◯ Unsure |
| I would use telemedicine again for future visits. | ◯ Strongly Agree<br>◯ Agree<br>◯ Neutral<br>◯ Disagree<br>◯ Strongly Disagree |
| I prefer in-person visits to telemedicine visits. | ◯ Strongly Agree<br>◯ Agree<br>◯ Neutral<br>◯ Disagree<br>◯ Strongly Disagree |
| If your primary doctor referred you to a specialist in the city, would you be more likely to see them if they could do a video visit instead of an in-person visit? | ◯ Strongly Agree<br>◯ Agree<br>◯ Neutral<br>◯ Disagree<br>◯ Strongly Disagree |

**Supplementary Table 2. Summary of Patient Cohort Primary Diagnosis Categories**

| Diagnosis Category | Primary Diagnosis | |
|---|---|---|
| **PD/Parkinsonism Movement Disorder 66 (48%)** | Parkinson's Disease | 59 (89) |
| | Secondary Parkinsonism | 3 (5) |
| | Atypical Parkinsonism | 4 (6) |
| **Non-PD/Parkinsonism Movement Disorder 42 (31%)** | Ataxia / Cerebellar Dysfunction | 10 (24) |
| | Dystonia | 4 (10) |
| | Chorea | 6 (14) |
| | Tremor | 10 (24) |
| | Functional Movement Disorder | 3 (7) |
| | Gait Disorder | 1 (2) |
| | Hereditary Spastic Paraplegia | 2 (5) |
| | Restless Legs Syndrome | 1 (2) |
| | Synkinesis | 1 (2) |
| | Tardive Dyskinesia | 1 (2) |
| | Abnormal Involuntary Movement | 1 (2) |
| | Neurogenetics Evaluation for NBIA | 1 (2) |
| **Non-Movement Neurological Disorder 29 (21%)** | Neck Pain | 2 (7) |
| | Awareness alteration | 1 (3) |
| | Dementia without behavioral disturbance | 1 (3) |
| | Facial Weakness | 1 (3) |
| | Meningioma | 1 (3) |
| | Headache | 14 (48) |
| | Myasthenia gravis | 1 (3) |
| | Neuropathy | 5 (17) |
| | Stuttering | 1 (3) |
| | Trigeminal Neuralgia | 1 (3) |
| | Winging of Scapula | 1 (3) |

Data reported as N (%) of total cohort.
Abbreviations: *PD, Parkinson's Disease*.

**Supplementary Table 3. Summary of Patient Impressions of Teleneurology Experience**

| Category | Question | Responses |
|---|---|---|
| **Ease of Joining Visit** | *It was easy to connect for the telemedicine visit.* | ***StrD***: 5 (3.65%)<br>***D***: 8 (5.84%)<br>***N***: 10 (7.30%)<br>***A***: 64 (46.72%)<br>***StrA***: 50 (36.50%) |
| **Patient Comfort** | *I felt comfortable asking my doctor questions about my diagnoses, treatment options, and research via video.* | ***StrD***: 0<br>***D***: 0<br>***N***: 6 (4.38%)<br>***A***: 31 (22.63%)<br>***StrA***: 100 (72.99%) |
| † **Visit Summary** | *I understood my plan of care and provider recommendations by the end of the visit.* | ***StrD***: 0<br>***D***: 0<br>***N***: 3 (2.19%)<br>***A***: 57 (41.61%)<br>***StrA***: 73 (53.28%) |
| **Quality of Care** | *I was satisfied with the care provided during my telemedicine visit.* | ***StrD***: 2 (1.46%)<br>***D***: 0<br>***N***: 9 (6.57%)<br>***A***: 50 (36.50%)<br>***StrA***: 76 (55.47%) |
| †† **Money Saved** | *I saved money using telemedicine compared to an in-person visit.* | ***StrD***: 1 (0.73%)<br>***D***: 6 (4.38%)<br>***N***: 38 (27.74%)<br>***A***: 48 (35.04%)<br>***StrA***: 44 (32.12%) |
| ††† **Time Saved** | *I saved time using telemedicine compared to an in-person visit.* | ***StrD***: 0<br>***D***: 1 (0.73%)<br>***N***: 11 (8.03%)<br>***A***: 51 (37.23%)<br>***StrA***: 74 (54.01%) |
| †††† **Future Care** | *I would use telemedicine again for future visits.* | ***StrD***: 0<br>***D***: 5 (6.76%)<br>***N***: 4 (5.41%)<br>***A***: 28 (37.84%)<br>***StrA***: 37 (50.00%) |
| ††††† **Model of Future Care** | *For future visits with your provider, would you like to use in-person visits, telemedicine visits, or a combination of the two?* | ***In-Person***: 15 (23.08%)<br>***Telemedicine***: 9 (13.85%)<br>***Combination***: 41 (63.08%) |

Unless specified, questions above were asked with a 5-point Likert-scale, with response options including Strongly Disagree (StrD), Disagree (D), Neutral (N), Agree (A), and Strongly Agree (StrA). Data reported as N (%) of total cohort.

† 4 (2.92%) patient responses missing.

†† Average dollar amount saved: $36.65, SD ± 43.58

††† Average self-reported time saved: 132 minutes, SD ± 152

†††† A subset of N=74 (54.01%) patients were asked this question.

††††† A subset of N=65 (47.44%) patients were asked this question.

**Supplementary Table 4. Patient Impressions About the Quality of the History Obtained During the Visit**

| Whole Cohort | Worse | Same | No. Responses |
|---|---|---|---|
| | 17 (12) | 120 (88) | 137 |
| **Gender** | | | |
| Male | 10 (16) | 54 (84) | 64 |
| Female | 7 (10) | 66 (88) | 73 |
| **Age Group (Years)** | | | |
| 18-29 | 2 (40) | 3 (60) | 5 |
| 30-49 | 2 (11) | 16 (89) | 18 |
| 50-69 | 8 (12) | 60 (88) | 68 |
| 70-89 | 5 (11) | 41 (89) | 46 |
| **Distance from Neurology Clinic (miles)** | | | |
| 0-19 | 9 (11) | 74 (89) | 83 |
| 20-39 | 5 (16) | 26 (84) | 31 |
| > 40 | 3 (13) | 20 (87) | 23 |
| **Visit Type** | | | |
| Existing Visit | 13 (13) | 88 (87) | 101 |
| New Visit | 4 (11) | 32 (89) | 36 |
| **Primary Diagnosis Category** | | | |
| PD/Parkinsonism Movement Disorder | 2 (7) | 27 (93) | 29 |
| Non-PD/Parkinsonism Movement Disorder | 8 (19) | 34 (81) | 42 |
| Non-Movement Neurological Disorder | 7 (11) | 59 (89) | 66 |

Data reported as N (%) of total cohort.
Abbreviations: *PD, Parkinson's Disease*.

**Supplementary Table 5. Patient Impressions About the Patient-Provider Relationship During Visit**

| Whole Cohort | Poor | Neutral/Fair | Good or Excellent | No. Responses |
|---|---|---|---|---|
| | 2 (1) | 9 (7) | 126 (92) | 137 |
| **Gender** | | | | |
| Male | 2 (3) | 5 (7) | 66 (90) | 73 |
| Female | 0 (0) | 4 (6) | 60 (94) | 64 |
| **Age Group (Years)** | | | | |
| 18-29 | 0 (0) | 1 (20) | 4 (80) | 5 |
| 30-49 | 0 (0) | 1 (6) | 17 (94) | 18 |
| 50-69 | 1 (1) | 4 (6) | 63 (93) | 68 |
| 70-89 | 1 (2) | 3 (7) | 42 (91) | 46 |
| **Distance from Neurology Clinic (miles)** | | | | |
| 0-19 | 2 (2) | 4 (5) | 77 (93) | 83 |
| 20-39 | 0 (0) | 2 (6) | 29 (94) | 31 |
| > 40 | 0 (0) | 3 (13) | 20 (87) | 23 |
| **Visit Type** | | | | |
| Existing Visit | 1 (1) | 8 (8) | 92 (91) | 101 |
| New Visit | 1 (3) | 1 (3) | 34 (94) | 36 |
| **Primary Diagnosis Category** | | | | |
| PD/Parkinsonism Movement Disorder | 0 (0) | 1 (3) | 28 (97) | 29 |
| Non-PD/Parkinsonism Movement Disorder | 2 (5) | 2 (5) | 38 (90) | 42 |
| Non-Movement Neurological Disorder | 0 (0) | 6 (9) | 60 (91) | 66 |

Data reported as N (%) of total cohort.
Abbreviations: *PD, Parkinson's Disease*.

**Supplementary Table 6. Patient Impressions About the Time Required During Visit**

| **Whole Cohort** | **Less** | **Same** | **More** | **No. Responses** |
|---|---|---|---|---|
| | 17 (19) | 64 (71) | 9 (10) | 90 |
| **Gender** | | | | |
| Male | 8 (17) | 32 (70) | 6 (13) | 46 |
| Female | 9 (20) | 32 (73) | 3 (7) | 44 |
| **Age Group (Years)** | | | | |
| 18-29 | 0 (0) | 2 (100) | 0 (0) | 2 |
| 30-49 | 1 (10) | 9 (90) | 0 (0) | 10 |
| 50-69 | 7 (16) | 33 (73) | 5 (11) | 45 |
| 70-89 | 9 (27) | 20 (61) | 4 (12) | 33 |
| **Distance from Neurology Clinic (miles)** | | | | |
| 0-19 | 11 (19) | 39 (68) | 7 (12) | 57 |
| 20-39 | 2 (11) | 15 (79) | 2 (11) | 19 |
| > 40 | 4 (29) | 10 (71) | 0 (0) | 14 |
| **Visit Type** | | | | |
| Existing Visit | 17 (19) | 64 (71) | 9 (10) | 90 |
| New Visit | 0 (0) | 0 (0) | 0 (0) | 0 |
| **Primary Diagnosis Category** | | | | |
| PD/Parkinsonism Movement Disorder | 1 (6) | 13 (76) | 3 (18) | 17 |
| Non-PD/Parkinsonism Movement Disorder | 6 (30) | 12 (60) | 2 (10) | 20 |
| Non-Movement Neurological Disorder | 10 (19) | 39 (74) | 4 (8) | 53 |

Data reported as N (%) of total cohort.
Abbreviations: *PD, Parkinson's Disease*.

**Supplementary Table 7. Patient Impressions About the Overall Quality of the Visit**

| Whole Cohort | Worse | Same or Better | No. Responses |
|---|---|---|---|
| | 25 (20) | 103 (80) | 128 |
| **Gender** | | | |
| Male | 14 (20) | 55 (80) | 69 |
| Female | 11 (19) | 48 (81) | 59 |
| **Age Group (Years)** | | | |
| 18-29 | 1 (20) | 4 (80) | 5 |
| 30-49 | 3 (18) | 14 (82) | 17 |
| 50-69 | 13 (20) | 51 (80) | 64 |
| 70-89 | 8 (19) | 34 (81) | 42 |
| **Distance from Neurology Clinic (miles)** | | | |
| 0-19 | 18 (23) | 60 (77) | 78 |
| 20-39 | 4 (14) | 24 (86) | 28 |
| > 40 | 3 (14) | 19 (86) | 22 |
| **Visit Type** | | | |
| Existing Visit | 19 (19) | 81 (81) | 100 |
| New Visit | 6 (21) | 22 (79) | 28 |
| **Primary Diagnosis Category** | | | |
| PD/Parkinsonism Movement Disorder | 4 (14) | 24 (86) | 28 |
| Non-PD/Parkinsonism Movement Disorder | 9 (24) | 29 (76) | 38 |
| Non-Movement Neurological Disorder | 12 (19) | 50 (81) | 62 |

Data reported as N (%) of total cohort.
Abbreviations: *PD, Parkinson's Disease*.
